## Supplementary material for "Immunoglobulin A as a key immunological molecular signature of post-COVID-19 conditions"

Graziele Fonseca de Sousa *et al.*

**This PDF file includes:**

Figs. S1 to S4

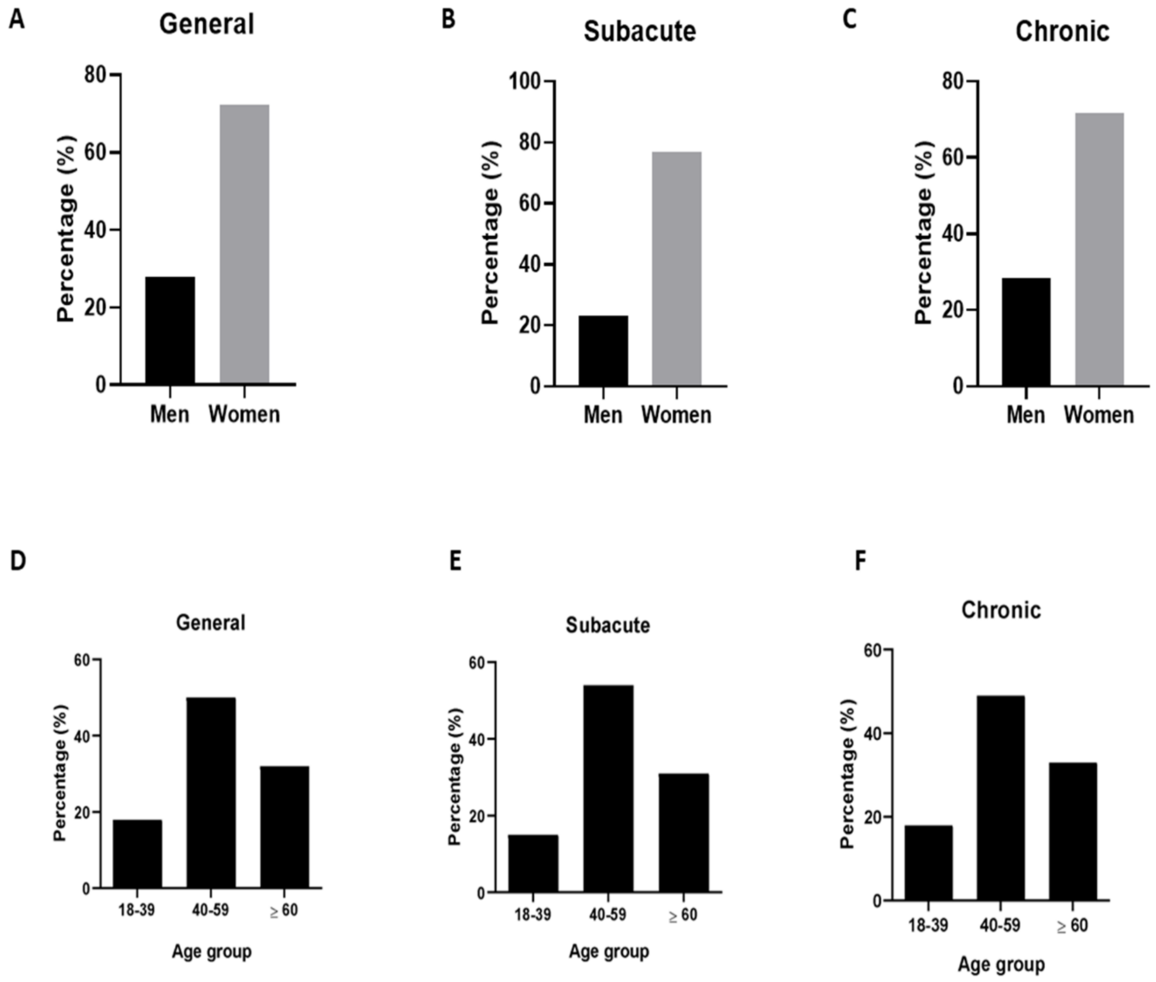


**Fig. S1. General data of the patients of the Centro de Acolhimento e Reabilitação Pós-COVID-19 (CARP).** Percentage of men and women who sought medical assistance at CARP (**A**), divided into subacute (**B**) and chronic (**C**) phases. Percentage of age groups that sought medical assistance at CARP (**D**), divided into subacute (**E**) and chronic (**F**) phases.

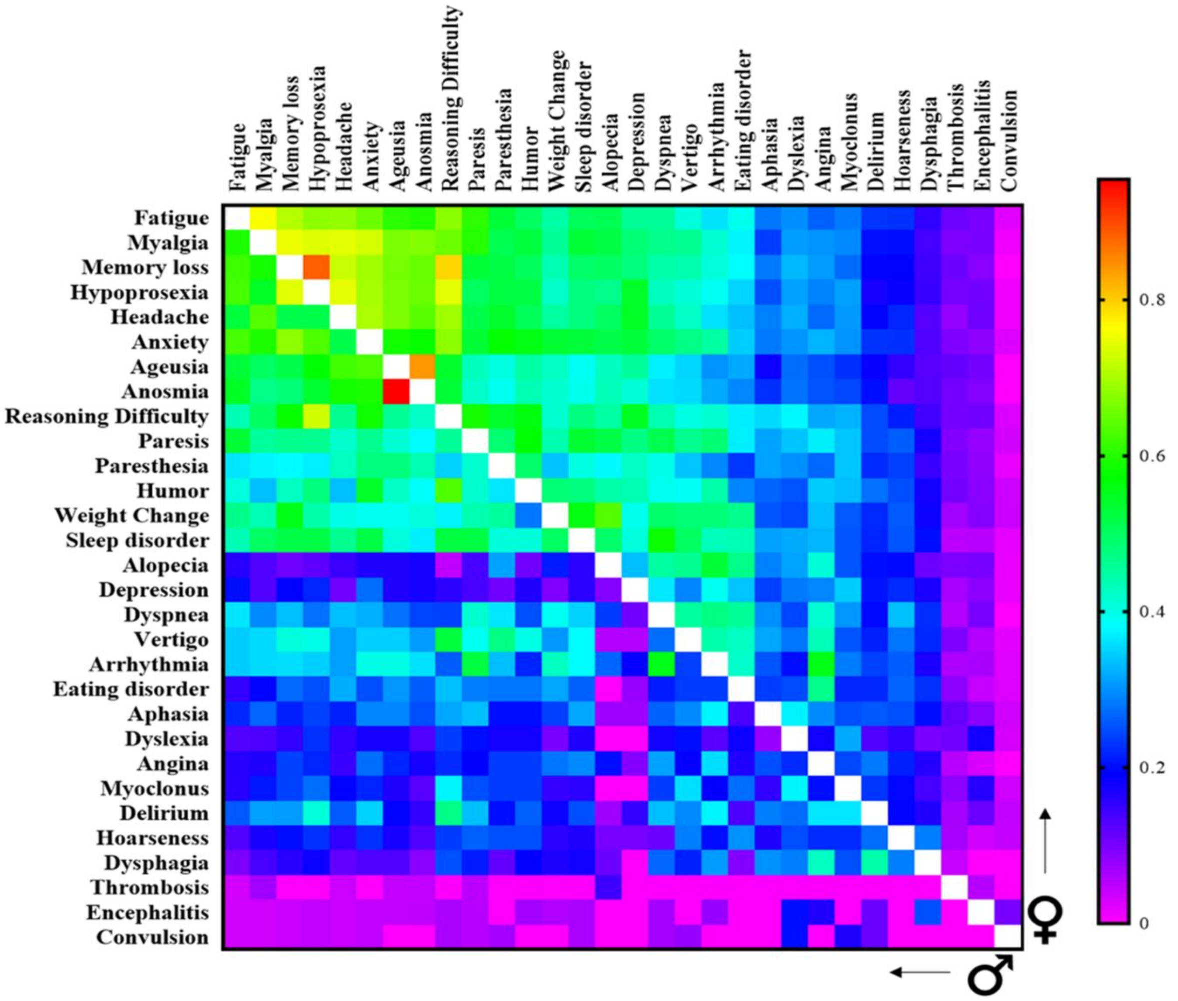


**Fig. S2. Heatmap showing co-occurrence levels (Jaccard similarity index) between pairs of post-COVID-19 condition symptoms in men and women.** The Jaccard index was measured based on the presence/absence of symptoms for each patient. Men are represented by the symbol (♂) and women by the symbol (♀). Increasing similarities are indicated by a warm color. Values equal to 1 are indicated by a blank square.

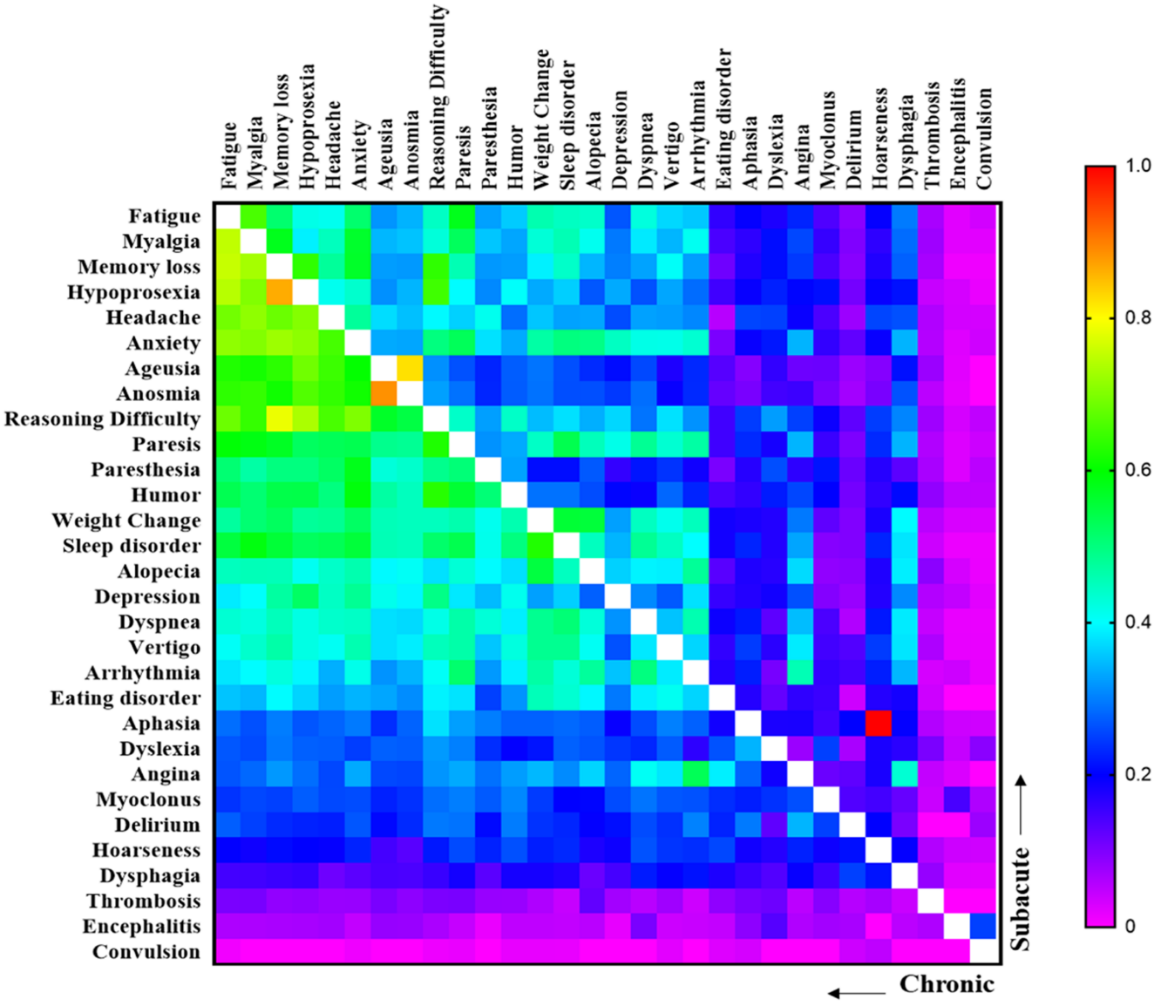


**Fig. S3. Heatmap showing co-occurrence levels (Jaccard similarity index) between pairs of post-COVID-19 condition symptoms in the subacute and chronic phases of PCC.** The Jaccard index was measured based on the presence/absence of symptoms based on the phase of the post-COVID-19 conditions. The co-occurrence of subacute phase symptoms is shown in the upper matrix, while the co-occurrence of chronic phase symptoms is shown in the lower matrix. Increasing similarities are indicated by a warm color. Values equal to 1 are indicated by a blank square.

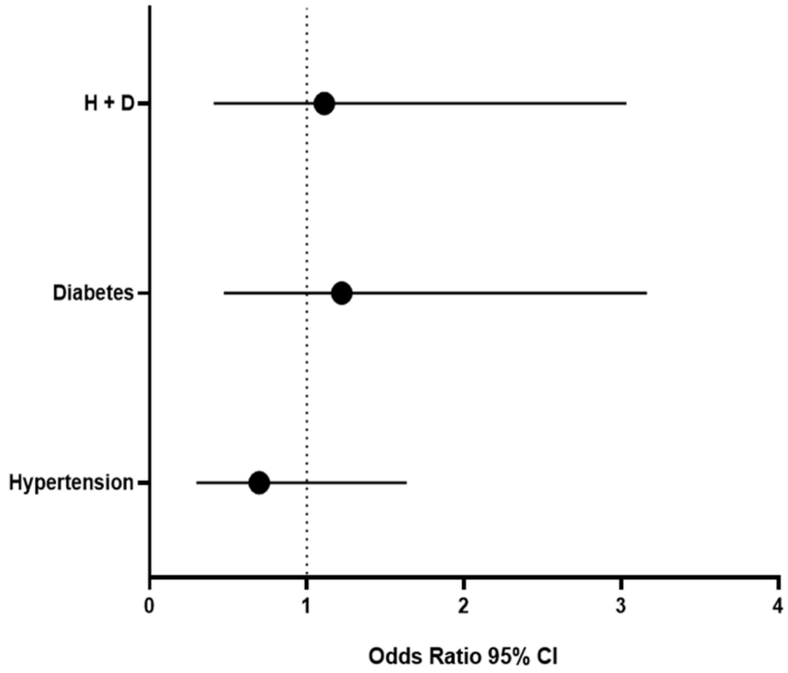


**Fig. S4. Comorbidities diabetes and hypertension as predisposing factors for the development of post-COVID-19 conditions (PCC).** Odds ratio with 95% confidence interval (CI) analysis of comorbidity domains for the development of chronic PCC symptoms. H+D represents patients with diabetes and hypertension comorbidities.
